# From Menarche to Menopause: Hormonal Influences on Functional Neurological Disorder

**DOI:** 10.64898/2026.07.16.26358260

**Authors:** David D. G. Palmer, Nicola Warren, Adam Morton, Alexander Lehn

**Author notes:** Corresponding Author Alex Lehn. Statement of Ethics This study protocol was reviewed and approved by Queensland Health’s Metro South Human Research Ethics Committee, approval number HREC/2023/MSH/00167. Data Availability Statement Data and analysis scripts are available through the Open Science Framework at https://doi.org/10.17605/OSF.IO/K3PYB.

## Abstract

**Background:** Functional neurological disorder (FND), one of the most common neurological conditions, affects women almost twice as frequently as men. The reasons for this are unknown, and there has been minimal research into how physiological and pathological features of women’s health interact with symptoms of FND.

**Methods:** We conducted an online survey assessing the effect of several aspects of women’s health with the severity of symptoms of FND.

**Results:** 484 people completed the survey. Among the 223 who had regular or fairly regular menstrual cycles, a strong difference across the menstrual cycle was seen, with symptoms at their best in the follicular phase, worsening in the luteal phase, and worst in the pre-menstrual period and the menses. This effect was not moderated by a proxy measure of pre-menstrual dysphoric disorder (PMDD).

Participants who were taking the combined oral contraceptive (COC, n=43) and progesterone-based contraception (n=80) were more likely to report symptom improvement from starting the medication than worsening. When compared to menstruating participants who were not taking the COC, participants taking the COC reported less worsening in their symptoms of FND in the luteal, pre-menstrual, and menstrual phases.

Of the 99 women who had passed menopause since developing FND, 76% reported worsening of their FND symptoms after menopause.

**Discussion:** This study demonstrates interactions between several aspects of women’s health and symptoms of FND. The observed pattern of symptom fluctuation across hormonal states suggests a potential modulatory role of oestrogen, warranting further targeted investigation.

## Introduction

Functional neurological disorder (FND) disproportionately affects women. Approximately 70% of people with the condition are of female sex, and this proportion is almost identical between functional movement disorder and functional seizure cohorts.^1,2^ Many possible reasons for the discrepancy in incidence between sexes have been proposed,^3–7^ however there is little evidence to support these, and the true reason for the difference remains unknown.

Proposed explanations for sex differences in the incidence of FND include both social and biological factors.^8^ Theories invoking social factors note higher rates of diagnostic labelling of symptoms as FND in women, as well as lower rates of investigation of symptoms,^7^ although these factors are less likely to account for differences in the present day, where diagnosis requires positive features of FND and the condition is no longer seen as a diagnosis of exclusion. The higher prevalence of psychological trauma—a recognised risk factor for FND—in women has also been hypothesized to contribute to sex-differences,^3–6^ and depression and anxiety, which also frequently co-occur with FND have higher prevalence in women.^9^ Proposed biological differences are less specific, however rates and symptoms of other neurological conditions are known to differ in females, and across changes in hormonal milieu for affected women, for example epilepsy, migraine, and Parkinson’s disease.^10^ Similar differences are seen in some psychiatric conditions which are recognised risk factors for FND, or which might have shared mechanisms.^9^ The prototype of these is premenstrual dysphoric disorder (PMDD), a condition marked by troublesome alterations in mood and neurocognitive function in the late luteal phase, which resolve with the onset of menses.^11^

Beyond the epidemiological data which describe the sex disparity of incidence, clinical observations suggest that, for some women, symptoms of FND may vary across their menstrual cycles. Some patients also describe changes in symptoms after passing menopause. Given the paucity of evidence on the subject, we aimed to measure the relationship between aspects of normal and ill-health related to female sex and variation in symptoms of FND.

While the overall aims of our study were descriptive, we also aimed to test one hypothesis. Given the association of both mood and physical symptoms with symptoms of FND,^3^ we hypothesized that variation of symptoms across the menstrual cycle might be accounted for by premenstrual dysphoric disorder, symptoms associated with menses, or both.

## Methods

### Questionnaire

We formulated a questionnaire designed to capture symptom severity and variation in relation to participants’ menstrual cycles, use of hormonal contraception, and experience of menopause. We also included the questions of the Daily Record of Severity of Problems (DRSP), an instrument used in the diagnosis of PMDD.^12^ The questionnaire was jointly designed by two neurologists with subspecialty expertise in FND (DP and AL), and was modified to improve clarity and acceptability based on input from four women with lived experience of FND. The questionnaire was administered online through SurveyMonkey, an online survey platform.^13^ A full transcript of the questionnaire is available as supplementary material.

### Participants and Data Collection

Participants were recruited to the study by following an online link. The link was disseminated by posts on the websites of FND Australia (fndaustralia.com.au), FND Hope (fndhope.org), and FND Australia Support Services (fndaus.org.au), by posts on social media, and through FND Research Connect (fnd-research.org), an online platform for recruitment of participants to FND studies. Enrolment time was not fixed, and was closed after three months, when the rate of survey responses had become minimal.

Recruitment was open to people aged 18 or older who were assigned female sex at birth and had been diagnosed with FND by a healthcare professional. Participants were screened by questions at the opening of the questionnaire, and the survey was automatically terminated if they did not meet the inclusion criteria.

### Data Analysis

Data were analysed using Python, with Bayesian regressions performed using Bambi,^14^ and k-means clustering performed using scikit-learn.^15^ Descriptive data are presented as means and standard deviations where data are normally distributed, and medians and interquartile ranges for non-normal data. Ordinal data from Likert scales were analysed using Bayesian ordinal regressions. Due to the difficulty of conveying ordinal data with summary statistics, results are presented graphically where possible. Where strengths of evidence from Bayesian models are presented in text, the categories of Jeffrey’s scale as refined by Lee and Wagenmakers^16^ are used.

To assess the possibility of PMDD in participants, we used the DRSP.^12^ The instrument divides questions into three parts: parts A and B assess emotional, and behavioural and physical symptoms, and part C assesses functional impairment due to these symptoms. Functional impairment is required to diagnose PMDD by the DSM-5 criteria,^11^ however, says little about the biological processes underlying the cyclical symptoms. We therefore assessed both the effect of symptoms of PMDD alone (Parts A and B), and of possible PMDD (including Part C), on symptoms of FND.

To test the hypothesis that there is a subgroup of women with FND who have cyclical variation in their symptoms of FND, we performed k-means clustering. The optimal number of clusters was chosen by inspection of an elbow plot and silhouette scores, and clustering was then performed using scikit-learn with the intention of performing between-groups comparisons of these clusters for both PMDD and period-related symptoms. On analysis, we found that the data did not support the hypothesis that there were discrete clusters of symptoms trajectories. We therefore performed an exploratory post hoc analysis using ordinal regression to compare symptom trajectories across the menstrual cycle of the group identified as having above-threshold scores on the DRSP to the group who did not.

## Results

### Participants and demographics

484 women completed the survey. The median age was 40 years, with quartiles at 30 and 49. The median time since first symptoms of FND was 5 years (quartiles 2 and 10 years), and the median time since diagnosis of FND was 1 year (quartiles 0 and 4 years). Participants endorsed a wide range of symptoms, encompassing all common presentations of FND. The symptoms reported by participants are presented in Supplementary Figure 1.

### Symptom Change Across the Menstrual Cycle

410 participants were menstruating. Of these, 223 rated their periods as ‘regular’, or ‘fairly regular’, and were included in analysis for this section. Of this group, 174 were taking no exogenous hormonal therapy, 21 were taking the COC, and 29 were using progesterone-based contraception. Response distributions at each phase of the menstrual cycle (excluding ‘Unsure’ responses) for these participants are presented in **Error! Reference source not found**. panel A.

The severity of symptoms of FND showed strong variation. The majority of participants rated their FND symptoms as ‘About Average’ in the follicular phase. This was also the only phase for which a meaningful proportion of participants (18%) reported their symptoms as being better than average. The majority response for the luteal phase was still ‘About average’, but almost as many participants (46%) rated their symptoms in one of the worse than average categories. In the pre-menstrual period, 84% of participants rated their symptoms as ‘Worse than average’, or ‘Much worse than average’. A similar proportion rated their symptoms in these categories during the menstrual period, but there was a small positive shift, with fewer participants rating their symptoms ‘Much worse than average’. Ordinal regression showed extremely strong evidence for differences between response patterns for all phases of the cycle.

Mean individual trajectories (change relative to the participant’s rating for the follicular phase) are presented in **Error! Reference source not found**. panel B, showing the same trend seen in overall numbers of responses per category.

With the hypothesis that there might be only a subgroup of women with FND who experience cyclical variation in their symptoms, we performed k-means cluster analysis on the data from this question. For values of k up to 9, no evidence for meaningful clustering was found by examination of an elbow plot or by comparison of silhouette scores. This was interpreted as evidence for lack of discrete clusters within the data, and cluster analysis was not continued.

**Figure 1.**
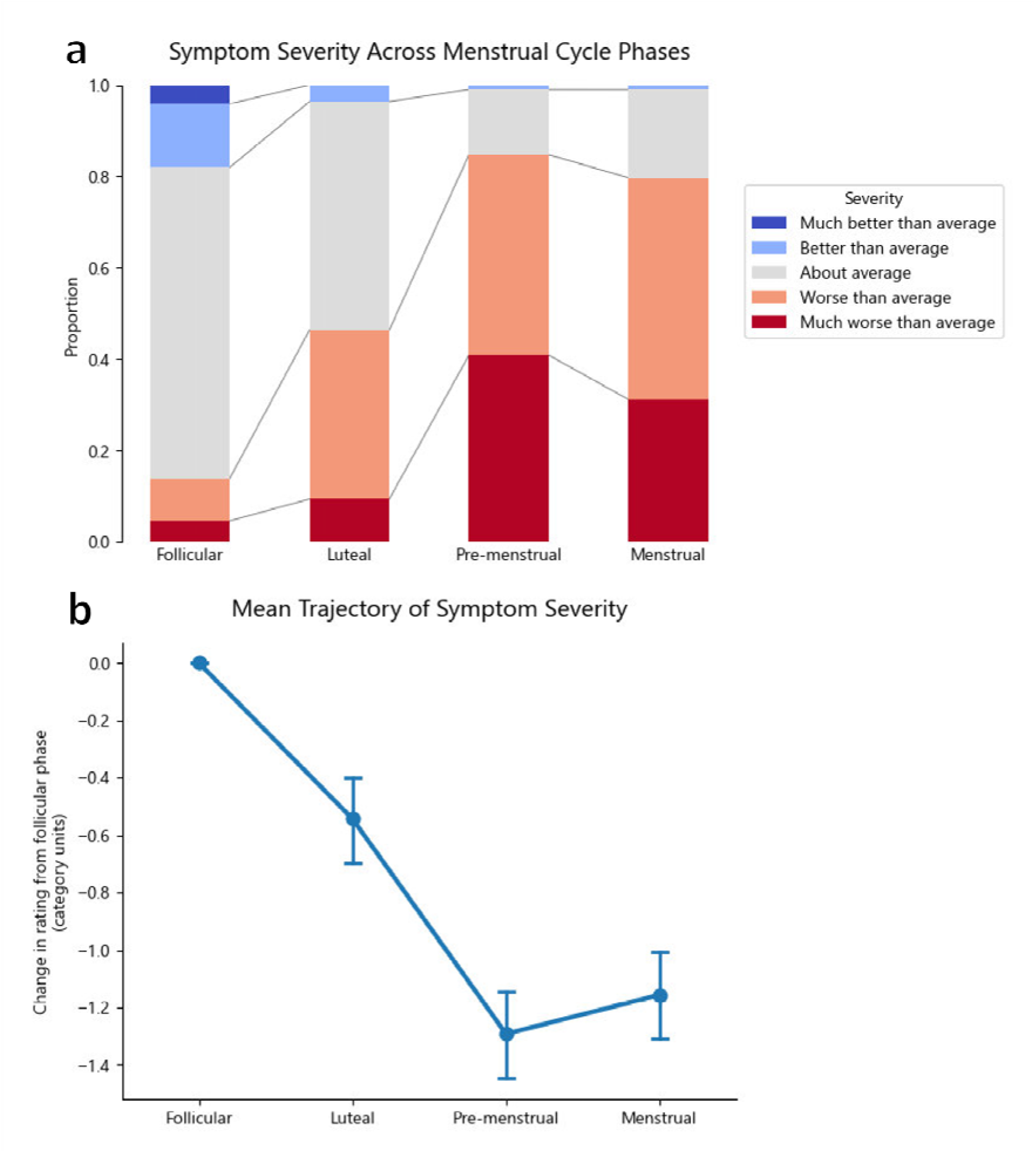
A: Aggregate symptom ratings by phase of the menstrual cycle. B: Mean trajectory (number of categories change from follicular phase rating). Points represent group mean and error bars represent bootstrapped 95% confidence intervals.

### Symptoms of PMDD

Of the 223 women with regular periods, 31.8% gave responses to the questions of the DRSP consistent with a diagnosis of PMDD, and 33.6% gave responses which would be consistent with having symptoms of PMDD but not fitting the diagnostic criteria due to lack of functional impairment.

With the failure of cluster analysis to detect discrete clusters in our data on FND symptom severity across the cycle, we performed a post-hoc analysis comparing the symptom trajectories of women who met the presumptive criteria for PMDD against those who did not. Symptom trajectories were calculated for each participant by calculating the difference between the symptom rating given for the follicular phase and each other phase. These trajectories were then compared between groups using ordinal regression. **Error! Reference source not found**. shows the outcome of this analysis. At baseline (follicular phase), women in the presumed PMDD group rated their symptoms as better than women without presumed PMDD, with standardised mean difference (SMD) of 0.4. Across all other phases, the ratings of the participants in the PMDD group showed greater worsening in their symptoms of FND compared to the group without presumed PMDD, with strong to extreme evidence for these differences.

**Figure 2.**
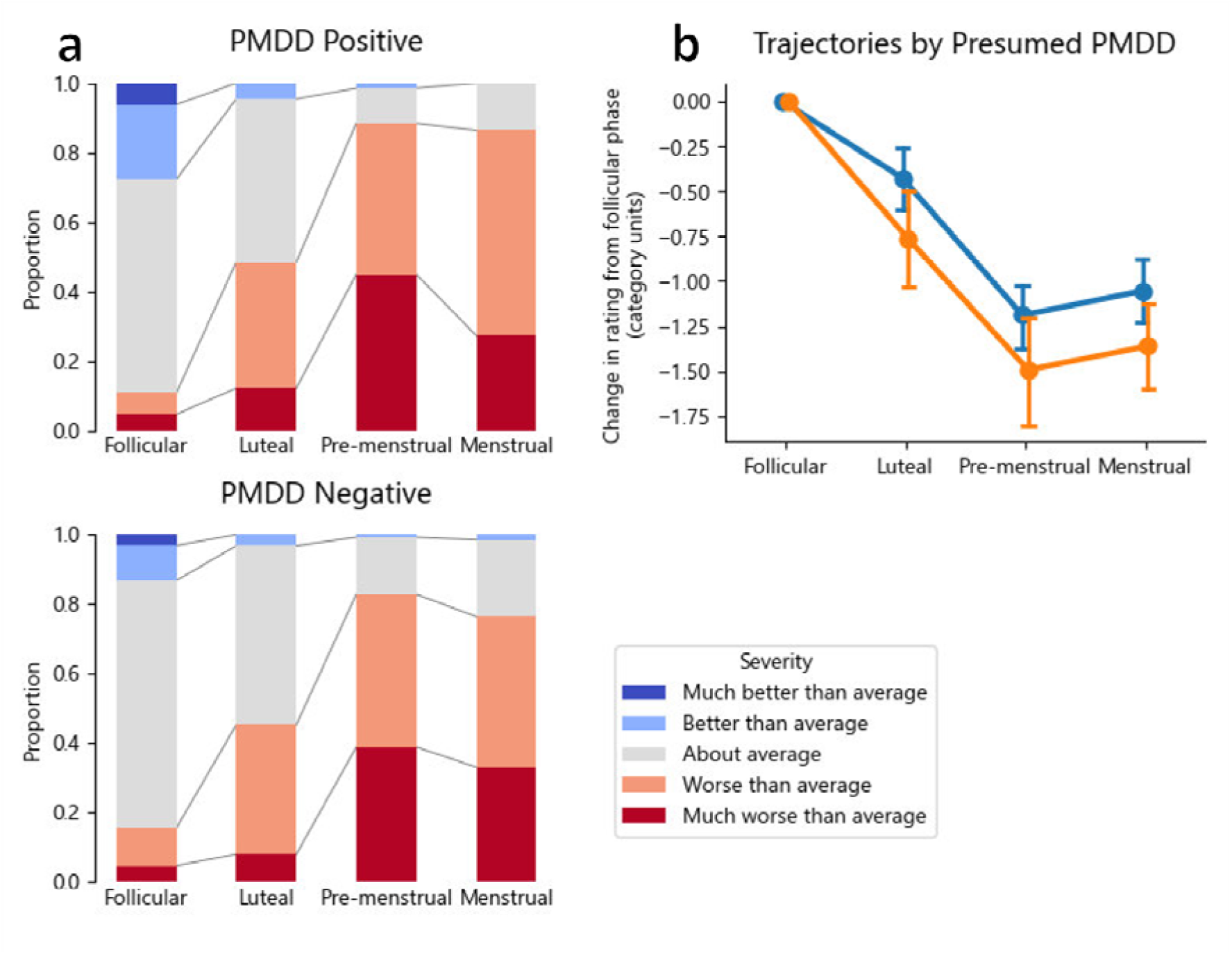
Aggregate symptom ratings by phase of the menstrual cycle. B: Mean trajectory (number of categories change from follicular phase rating). Points represent group mean and error bars represent bootstrapped 95% confidence intervals.

### The Effect of Hormonal Contraception

43 respondents were taking a combined oral contraceptive (COC), and 80 were using progesterone-based contraception. Responses to a question asking about the impact of starting contraception on symptoms of FND are presented in **Error! Reference source not found**. panel A. 8 respondents in the combined oral contraceptive group and 22 in the progesterone-based contraception group answered ‘Unsure’ to this question, and are excluded from the plot. In both groups, the modal response was ‘No change’. 26% of the COC group and 25% of the progesterone-based group reported FND symptom improvement on starting contraception, while 8% of the COC group and 15% reported symptom worsening.

**Error! Reference source not found**. panels B and C show the symptom trajectories and ratings of FND symptoms across the cycle for the 223 participants with regular periods comparing those taking and not taking the COC or progesterone-based contraception. In the COC group, categorical regression showed strong evidence for lesser symptom severity in the luteal phase, and extreme evidence for lesser symptom severity in the pre-menstrual phase for participants taking the COC. No strong evidence was found for any interaction between progesterone based contraceptive use and symptom severity at any phase of the cycle. Note that for consistency we have continued to use the terms follicular phase and luteal phase to refer to the first and second portions of the cycle in all participants, however these terms do not imply the same endocrine states in people taking the COC.

**Figure 3.**
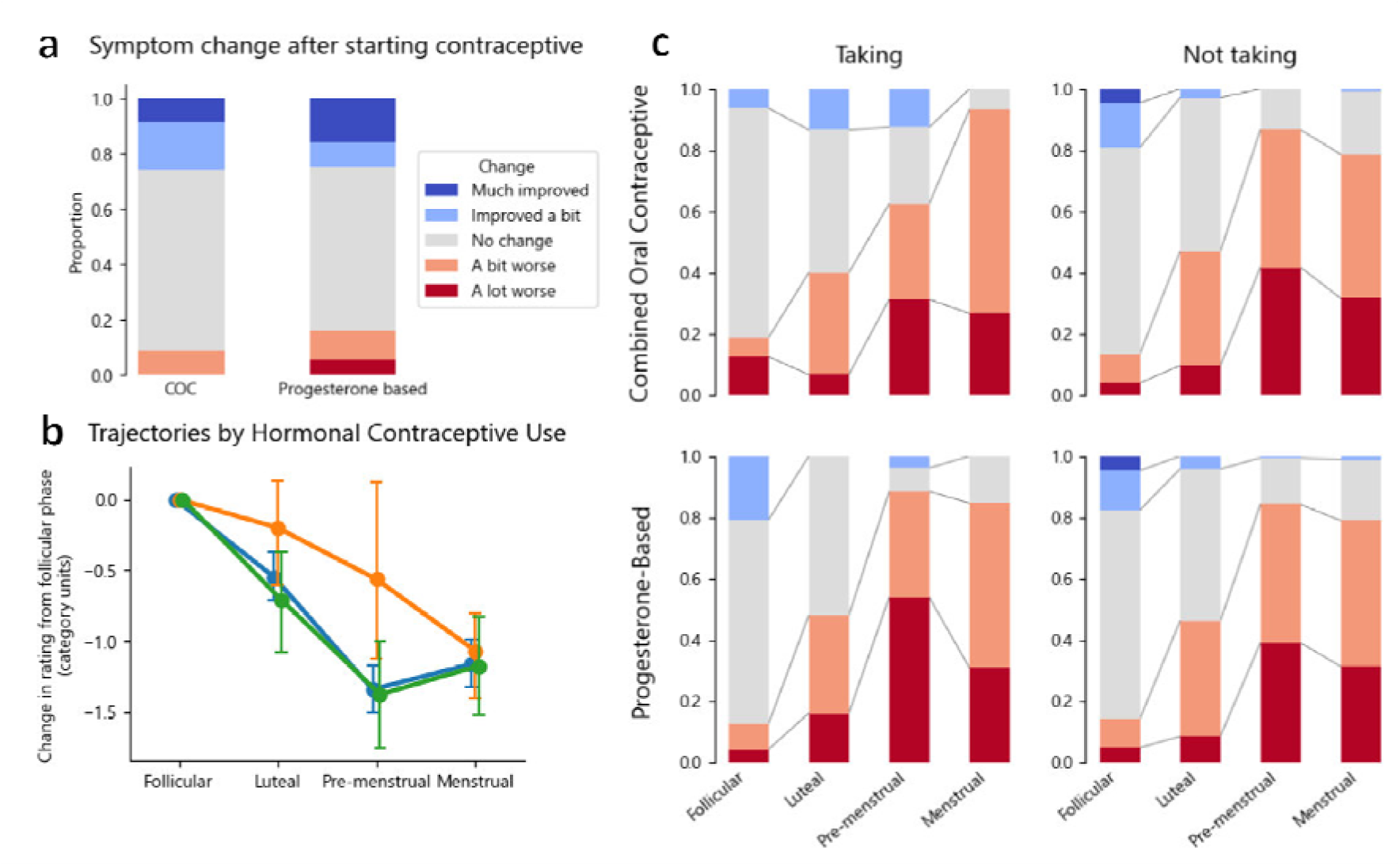
Effect of hormonal contraceptives on symptoms of FND. A: Change in symptoms after starting medication. B: Comparison of symptom trajectory (number of categories changed from follicular phase) between participants taking no hormonal contraception (blue), using progesterone-based contraception (green), and using the COC (orange). C: Aggregate symptom ratings by phase stratified by type, and use of hormonal contraception.

### The Effect of Menopause

99 participants had FND before menopause and had since passed menopause. Their responses for change in FND symptoms at menopause are presented in **Error! Reference source not found**.. The majority of participants (76%) reported that their symptoms of FND worsened after menopause, with only 4% reporting symptom improvement.

**Figure 4.**
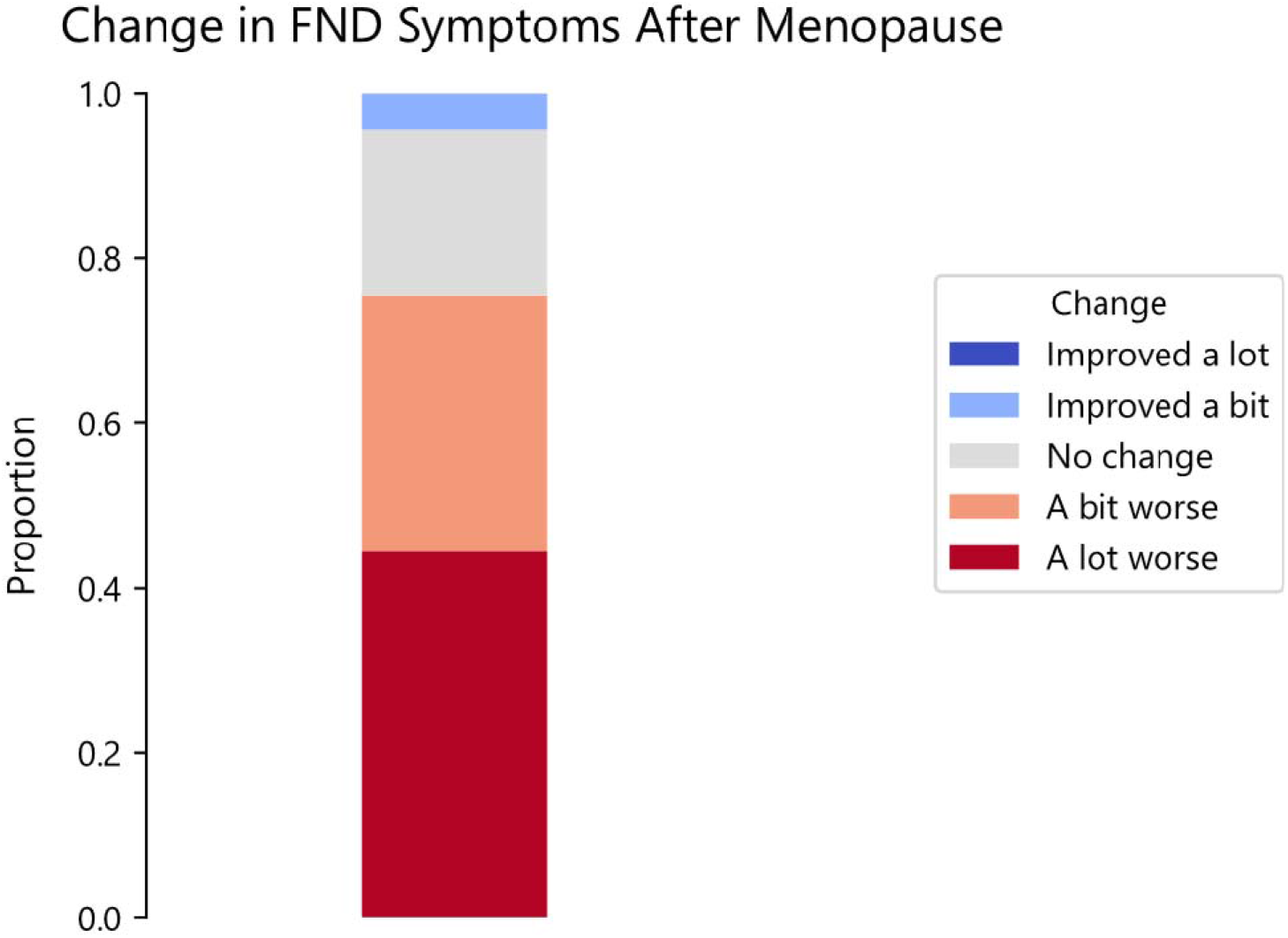
Change in severity of FND symptoms after menopause.

## Discussion

In this study, we have described the relationship between physiological and pathological experiences of women’s health and symptoms of FND across a large (n=484) cohort of women with FND. By examining such a large cohort, we have shown empirical evidence for relationships which have been suggested anecdotally, as well as identifying patterns have not been previously suggested.

For participants who were menstruating and had sufficiently regular periods to facilitate reporting of symptoms at different menstrual phases, we found marked systematic variation in symptoms of FND across the cycle, with symptoms at their mildest in the follicular phase, worse in the luteal phase, and at their worst across the pre-menstrual phase and the menses.

By using the questions of the DRSP, we were able to estimate which participants were likely to be affected by PMDD. Somewhat surprisingly, this presumptively identified PMDD did not seem to moderate the relationship between menstrual phase and symptom severity, although small effects were seen for greater improvement of symptoms in the follicular phase for the group with PMDD and greater worsening compared to the follicular phase across other phases of the cycle.

When comparing the group taking combined oral contraceptives to those not taking them, we saw reduced worsening of symptoms after the follicular phase. This, in concert with the findings described above, is suggestive for a possible role of reproductive hormones in moderating the symptoms of FND. Based on the trajectory of symptom severity across the menstrual cycle, one might hypothesise that—of the possible hormonal contributors—it is most likely that oestrogen effects symptom severity, given oestrogen levels are highest in the follicular phase,^17^ and that oestrogen supplementation with the COC is associated with reduced worsening in other phases of the cycle, when oestrogen levels are generally lower. While progesterone concentrations are high in the luteal phase when symptoms begin to worsen, both progesterone and oestrogen concentrations are low during the menses, and we saw no improvement in symptoms at that time.

Based on our data, it is possible either that falling levels of oestrogen (or oestrogen withdrawal with the placebo phase in in COC) precipitate worsening of symptoms, or that higher levels of oestrogen improve symptoms of FND. However, analogy to other conditions known to vary with fluctuating oestrogen concentrations, such as catamenial migraine^18^ and pre-menstrual exacerbation of symptoms of psychiatric conditions,^19^ suggest the former to be more probable.

Our ability to make inferences about the effect of supplemental progestogens is limited by the fact that we gathered data only on the use of these medications and not on the type, meaning we are unable to know which patients had meaningful systemic exposure to progestogens from their contraception. Since different forms of progesterone-based contraception have significantly different magnitudes of effect on oestrogen levels, further research examining the effects of different forms of progesterone-based contraception, different forms of COC, and different methods of administration of the COC (for instance, taking vs skipping placebos) on symptoms of FND is likely to be informative.

Three quarters of participants who had passed menopause since developing FND reported that their symptoms worsened after menopause. Our assessment of menopausal effects was limited, relying on a single item addressing change in symptoms across menopause, and therefore multiple explanations for this finding are possible. FND is frequently worsened by symptoms of other conditions, so the symptoms associated with the climacteric might plausibly aggravate symptoms of FND. Psychosocial factors also frequently change across the menopause,^20^ and might secondarily affect symptoms of FND. However, while these factors should not be discounted, if our suggestion of a role for oestrogen in modulating symptoms of FND is correct, the reduction in circulating oestrogen through the climacteric and menopause might suggest a more directly biological basis for this phenomenon.

Our results suggest potential new avenues of investigation for treating symptoms of FND. If falls in oestrogen or progesterone concentration are the precipitant for worsening of FND symptoms, strategies used in other conditions where catamenial variations in symptoms occur, such as menstrual migraine, may be worthy of study. Approaches supported by evidence for such conditions include longer courses of continuous hormonal replacement with less frequent placebo intervals,^21^ shorter placebo intervals with monthly COC preparations,^22^ administration of supplemental oestrogen in the pre-menstrual and menstrual periods,^23,24^ and the use of gonadotropin-releasing hormone (GnRH) to induce hypogonadotropic hypogonadism with continuous “add-back” oestrogen and progesterone replacement.^25,26^ Since ours is purely observational data, any potential treatments should be tested in a clinical trial before recommendations are made to patients. Additionally, studies examining the specific symptoms of FND that worsen or improve at different times during the menstrual cycle may be valuable in targeting future interventions.

As a cross-sectional study using survey responses, and with breadth limiting the opportunity for detail on many topics, our study does have important limitations. Questions about cyclical symptoms are dependent on shared understanding of timing, and it is possible that different respondents interpreted the descriptions of the timepoints of the menstrual cycle which we asked about differently. In addition, the reliance on retrospective report of symptom severity means that our results are vulnerable to recall bias based both on the general difficulty of recalling the timing of symptom fluctuations, and the possibility that recalled fluctuations were anchored to events in the menstrual cycle which are more salient in memory.

Our assessment for pre-menstrual dysphoric disorder is likely to be less accurate than a clinical diagnosis. Retrospective reports of PMDD symptoms are known to correlate poorly with prospective daily ratings,^12,27^ and therefore our use of the DRSP as a retrospective report tool should be regarded as giving at best a rough indication as to which participants may have PMDD. Furthermore, some of the physical and constitutional symptoms of PMDD which are assessed for in the DRSP are also symptoms of FND. It is therefore plausible that worsening of symptoms of FND in the pre-menstrual period could cause higher scores on the DRSP, limiting criterion validity of the instrument in this cohort. It is notable that the proportion of participants who we identified as having possible PMDD is markedly higher than the proportion of the general population who have PMDD,^28^ adding weight to the suggestion that our appraisal of participants as having possible PMDD was non-specific.

Finally, while not a methodological issue per se, it should be noted that this study focused only on experiences related to biological female sex, and not to female gender. Many experiences of people of female gender differ from those who are not of female gender for reasons which are not biological, but which might still be relevant to the development and experience of FND. These are likely to be fertile ground for future study.

In summary, this study has demonstrated large changes in severity of symptoms of FND in relation to several phenomena related to female sex, including differences across the menstrual cycle, the menopause, and in relation to use of the COC. Given FND’s greater tendency to affect women than men, further investigation of the mechanisms and potential therapeutic implications of these trends is an important next step in understanding and treating this condition.

## Data Availability

Data and analysis scripts are available through the Open Science Framework at https://doi.org/10.17605/OSF.IO/K3PYB.

## Statement of Ethics

This study protocol was reviewed and approved by Queensland Health’s Metro South Human Research Ethics Committee, approval number HREC/2023/MSH/00167.

## Author Contributions

DDGP: Conceptualization, Formal Analysis, Investigation, Methodology, Visualization, Writing – original draft, Writing – review & editing; NW: Writing – review & editing; AM: Writing – review & editing; AL: Conceptualization, Investigation, Methodology, Writing – review & editing.

