## Supplementary material for "From Menarche to Menopause: Hormonal Influences on Functional Neurological Disorder": Supplemantary material

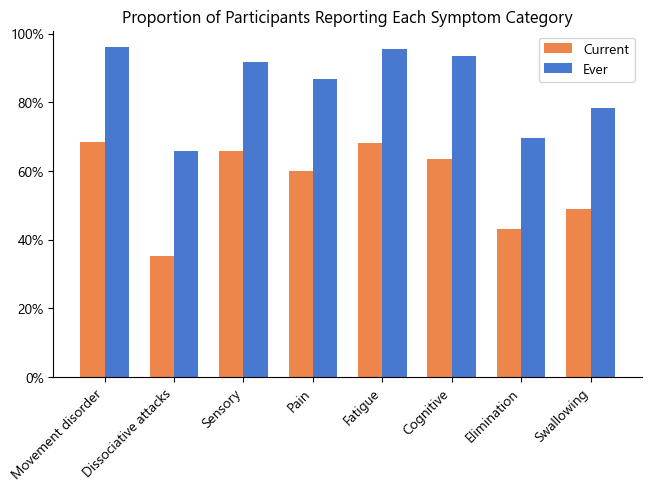

**Supplementary figure 1:** Proportion of participants reporting having experienced symptoms from each category in the last month (orange) or at any time in life (blue).
